# Suppression of COVID-19 infection by isolation time control based on the SIR model and an analogy from nuclear fusion research

**DOI:** 10.1101/2020.09.18.20197723

**Authors:** Osamu Mitarai, Nagato Yanagi

## Abstract

The coronavirus disease 2019 (COVID-19) has been damaging our daily life after declaration of pandemic. Therefore, we have started studying on the characteristics of Susceptible-Infectious-Recovered (SIR) model to know about the truth of infectious disease and our future.

After detailed studies on the characteristics of the SIR model for the various parameter dependencies with respect to such as the outing restriction (lockdown) ratio and vaccination rate, we have finally noticed that the second term (isolation term) in the differential equation of the number of the infected is quite similar to the “helium ash particle loss term” in deuterium-tritium (D-T) nuclear fusion. Based on this analogy, we have found that isolation of the infected is not actively controlled in the SIR model. Then we introduce the isolation time control parameter q and have studied its effect on this pandemic. Required isolation time to terminate the COVID-19 can be estimated by this proposed method.

To show this isolation control effect, we choose Tokyo for the model calculation because of high population density. We determine the reproduction number and the isolation ratio in the initial uncontrolled phase, and then the future number of the infected is estimated under various conditions. If the confirmed case can be isolated in 3∼8 days by widely performed testing, this pandemic could be suppressed without awaiting vaccination. If the mild outing restriction and vaccination are taken together, the isolation control time can be longer. We consider this isolation time control might be the only solution to overcome the pandemic when vaccine is not available.

## 1. Introduction

Since the Wuhan pneumonia in this Jan. 20 in China, was reported, we have been studying the COVID-19 infection using SIR model. Then, we have noticed the second equation (the quarantine term) used in the SIR mode is quite similar to the ^4^He ash particle balance equation in the nuclear fusion research [1]. By this analogy, we have found that the time in the isolation term can be interpreted as the natural isolation, which means that the isolation is not conducted in a controlled manner in a traditional SIR model. This fact inspired us how to suppress this infectious disease. We show that it is possible to calculate the required isolation time (from infection to isolation) to suppress the COVID-19.

In this paper we have chosen Tokyo with large population where might be suitable to apply the SIR model. The future number of the infected is estimated with various conditions with the outing restriction ratio and isolation time control parameters. When the infected people are identified by massive testing and then quickly isolated from the society, the infectious disease could be terminated.

## 2. Estimation of the future number of infection in Tokyo based on SIR model

In this paper we have used the traditional SIR model [2][3]. In this model we interpret the isolation term using the analogy from the nuclear fusion research, providing the new insight into this field. SIR model equations are given by

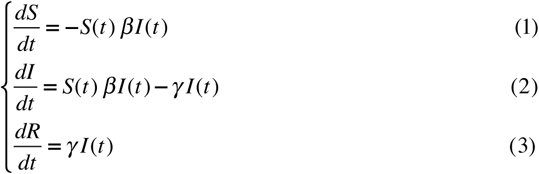

where *S* is the susceptible fraction, *I* is the infectious fraction and *R* is the recovered fraction which is the sum of the death toll and recovered. β is the infection rate per day and γ is the isolation ratio or the recovered rate per day.

The second term in the right-hand side of Eq. (2) is called the isolation term or quarantine term, and it can be also expressed by

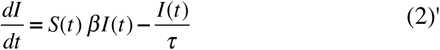

where τ has the dimension of time expressing the averaged lifetime of the infectious disease. This is also interpreted that an infected person is finally removed from the society by recovery or death after the time period of τ [2]. This isolation term in Eq. (2) is quite similar to the “^4^He ash particle loss term” used in a “nuclear fusion research” [1]. This loss time is called the “particle confinement time” in a nuclear fusion, and expresses the average time for ^4^He particle to escape from the confined plasma. This analogy inspired us to consider how to terminate the COVID-19 pandemic. Detailed explanations on this analogy will be described in the Appendix. If the isolation can be quickly conducted within a short time, isolation term is increased, and then the number of infected people would be reduced. This is because the contact time with infected people can be shortened.

In this SIR model, the total population is constant as N(t) = S(t)+I(t)+R(t) = constant by adding Eqs.(1)-(3). Therefore, we have calculated the number of the infected in Tokyo because of a constant population in a short time. To solve the SIR model, the initial value is set to S(0)=1, R(0)=0 and I(0)=given. We also note that this traditional SIR model assumes the permanent immunity.

Using the effective reproduction number defined by R_eff_= βS(t)/γ, Eq.(2) can be written as dI/dt = γ (R_eff_ -1)I. As the basic reproduction number R_o_= β/γ is for the initial value of S(0)=1, R_eff_=S(t)R_o_ holds. We see that the number of the infected people increases for R_eff_>1 and decreases for R_eff_<1, where R_eff_ expresses the positive or negative value corresponding to the derivative dI/dt. R_eff_ only shows the tendency at the present time slice.

### 2.1. First phase

We divided the calculated time ranges into the first phase and second phase in the following. The first phase starts on May 1, 2020 in this study and may not be affected by any governmental control measures. As the Day 48 from the start (April 17, 2020) provides the largest number of the infected per day, the two days later (Day 50) was taken as the first day of the second phase.

The first phase is given by the following traditional SIR model expressed by the basic reproduction number R_o_ as

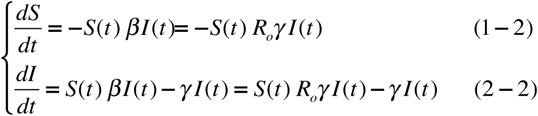

As Eq. (3) is the same as the second term in Eq.(2-2) with the inverse sign, we do not write down it in the following for simplicity. Here the only one infected person is assumed as the initial value I(0)= 1/14,000,000=7.14286 x10^−8^ and S(0)=1 for the population of Tokyo 14,000,000 as given in Table 1. We calculate the fraction of *S, I* and *R*, and multiply the population to obtain the final value. Above simultaneous equation has been solved by Mathematica (wolfram research).

**Table 1.**
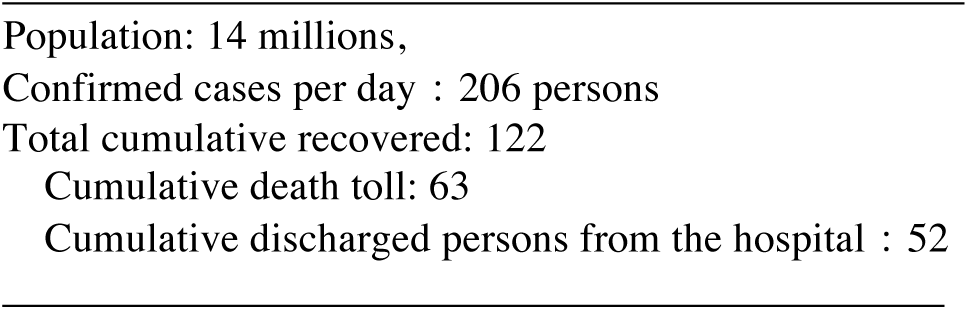
Data of COVID-19 patients in Tokyo (April 17, 2020) [4]

Although we need parameters β, γ and then the basic reproduction number R_o_, it is difficult to determine γ by Eq.(3) because data is not enough and not smooth in this case. As the basic reproduction number is reported to be 2 to 3 by recent research [5], we have used this range of value. As the main purpose of this paper is to estimate the isolation time and not to analyze the present situation in detail, the recovery time is most important. Therefore, we surveyed the recovery time using the cumulative number of the confirmed case and discharged case from the hospital as shown in Fig. 1-(a). The time difference between two curves is considered to be nearly the recovery time, which is less than 20 days.

**Fig. 1.**
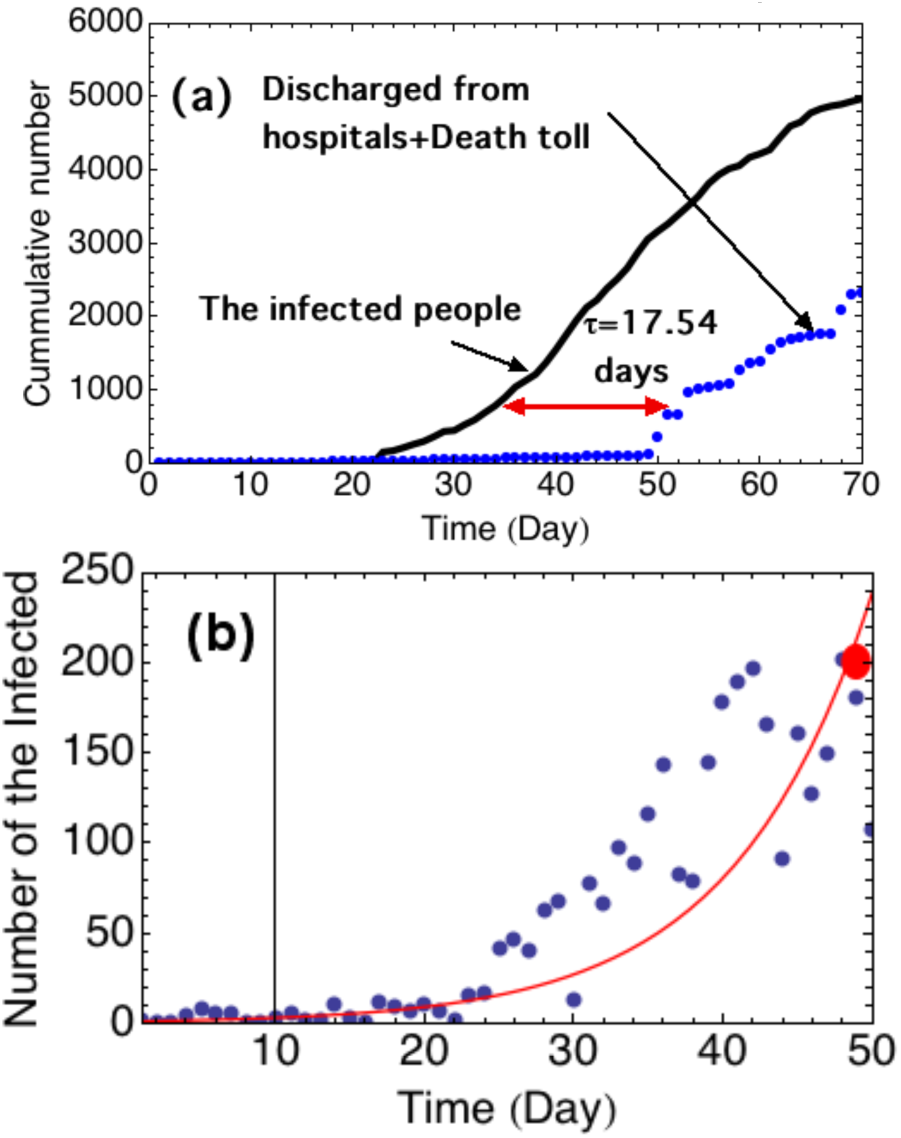
(a) Cumulative number of the infected, and the discharged from the hospitals and death toll as a function of time [4]. (b) The number of the infected people calculated by SIR model (Red line) and actual data (blue dots) in the non-controlled first phase. β = 0.1653, γ = 0.057 and R_o_ = 2.9 are determined by this fitting to the data base [7].

As Eq. (2-2) provides the number of the infected, γ and R_*o*_ are chosen by the trial and error to fit with the infection data as shown in Fig.1-(b). Then we have employed R_o_=2.9 as given by Liu et al. [6], and γ=0.057 corresponding to the recovery time τ=17.54 days less than 20 days, which provide the coefficients β = 0.1653.

### 2.2. The second phase after the outing restriction

In this work the second phase has started after 12 days from the state of emergencies in Japan (April 7). After Day 50 we introduce the outing restriction ratio c = 80 % in the SIR model, which means that 20% of the total population can move freely outside and 80% are staying home. This calculation can be done by employing the basic reproduction number R_o_ multiplied by (1-c) [8]. This means that the number of the susceptible people is reduced by (1-c), namely S(t)(1-c), and the number of non-susceptible people of S(t)c is staying home without infection. With the outing restriction ratio c, Eqs. (1) and (2) can be written as

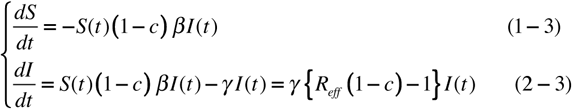

where R_eff_=βS(t)/γ=S(t)R_o_, The calculated results are shown in Fig. 2 for various outing restriction ratios c. The initial values in the second phase are taken from the last one in the first phase. A small number of c is not effective at all to reduce infection. A large number, such as c=0.8, can reduce the number of the infected substantially, but it takes two months (June 18, 2020) to reduce down to 50 persons per day, and four months down to 10 persons per day.

**Fig. 2.**
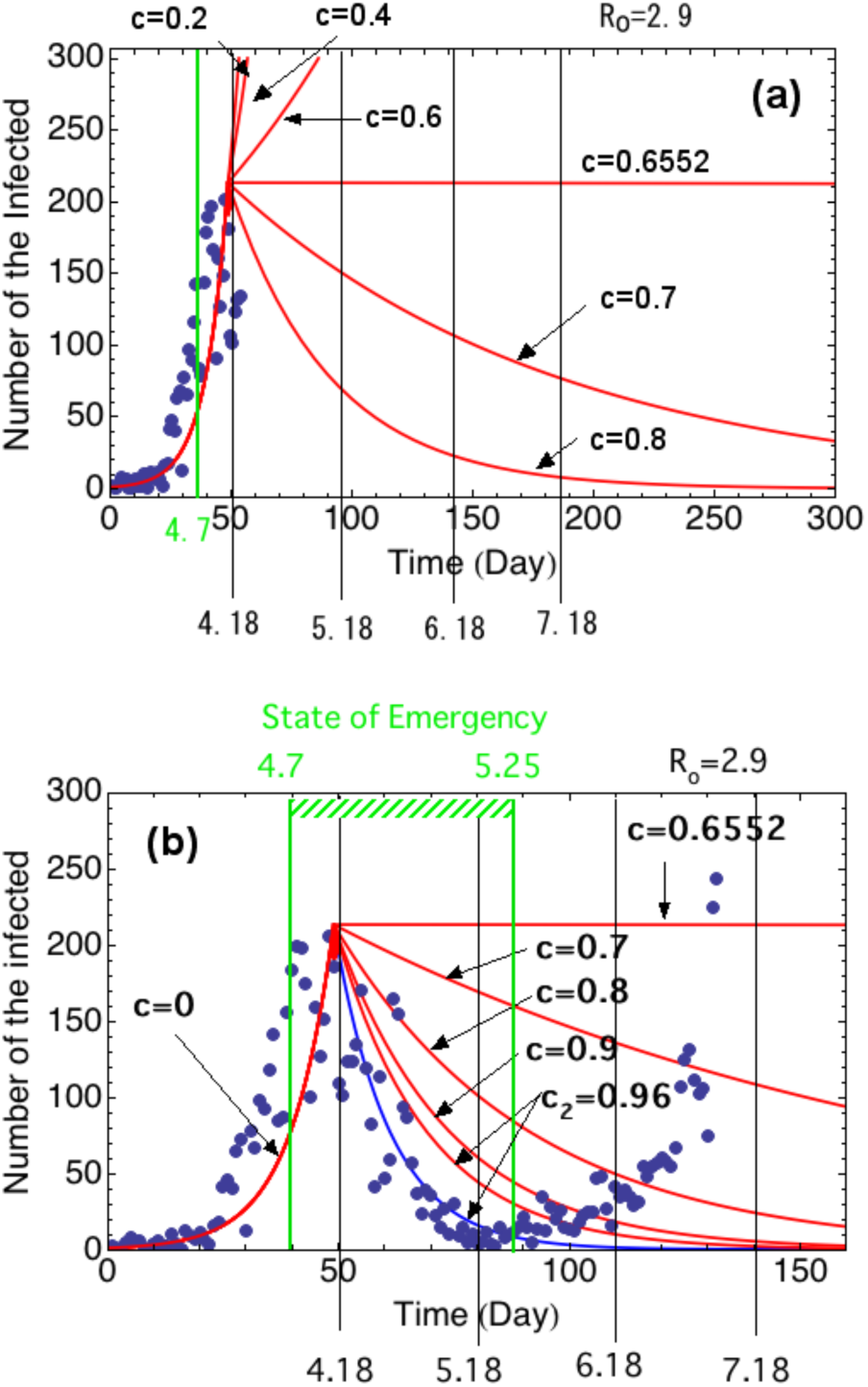
Temporal evolution of the number of the infected for various outing restriction ratios in the second phase during and after the state of emergency. Green line at April 7 shows the state of emergency. (a) Calculated at April 20, 2020, and (b) calculated at July 10, 2020. Note that red lines show the cases of the isolation ratio of γ=0.057 (R_o_=2.9) and a blue line γ=0.09 (R_o_=1.83).

On the other hand, if c=0.7 is taken it will take 200 days to reduce down to 50 persons per day. If c=0.65 is taken, the number of infected per day is not reduced and constant. This value can be derived by the condition of R_o_(1-c) =1 obtained by dI/dt=0 and S(t)∼1 in Eq. (2-3), providing c=1-1/R_o_=1-1/2.9 =0.655. When c=0.6 is chosen, the number of the infected is increasing as shown in Fig.2-(a) We should note that the variation of the outing restriction leads to the large difference in the number of the infected.

We added the data until July 9, 2020 to compare it with the actual situation as shown in Fig.2-(b). At the first glance, the state of emergency was so effective to reduce the infection. The actual number of the infected is much smaller than the curve of c=0.8 and 0.9 near the end of the state of emergency. This is because we assume the outing restriction is imposed on only the susceptible people and the infected patients are free to move. As the outing of the infected patients is also limited, the first term in Eq.(2-3) should be *S(t)(1-c)*^*2*^*βI(t)* for the assumption of the same outing restriction ratio. Therefore, the outing restriction of c=0.8 actually corresponds to c_2_=1-(1-0.8)^2^= 0.96 if we transform the form (1-c)^2^ to (1-c_2_). Although larger value of c=0.8 is proposed by committee, larger value of c_2_=0.96 was achieved in an actual situation. This estimation can be justified by the two-body collision theory as also used in nuclear fusion research [1], corresponding to the first term in Eq.(A-1) in Appendix. But the red curve with c_2_=0.96 is still higher than the actual values. Therefore, we increased the isolation ratio from γ=0.057 (red line) to γ=0.09 (blue line), providing the reasonable fit. The larger isolation ratio could be justified by active isolation conducted during this period.

It is criticized that polymerase chain reaction (PCR) testing is not fully conducted to find the COVID-19 patients in Japan. Therefore, more infected patients are expected [9]. However, as the purpose of this work is to see the future tendency and to study the isolation effect, we use the presently available data without discussion on this problem.

### 2.3. The third phase with the second wave

Even after the first wave is over, some portion of infected people still exit. Therefore, the infected patient would be increased again as long as outing restriction is lifted up. We predict the number of the infected as a function of the outing restriction ratio in the third phase that started after 100 days.

In this calculation, the outing restriction ratio in the second phase (50 to 100 days) is assumed as c=0.9. In Fig. 3-(a), the numbers of the infected in the third phase are shown for c=0.5 to 0.8. When the outing restriction is set to c=0.5, it increases again up to 200 infected people per day in September. For c=0.6, it increases to the same level by the end of 2020.

**Fig. 3.**
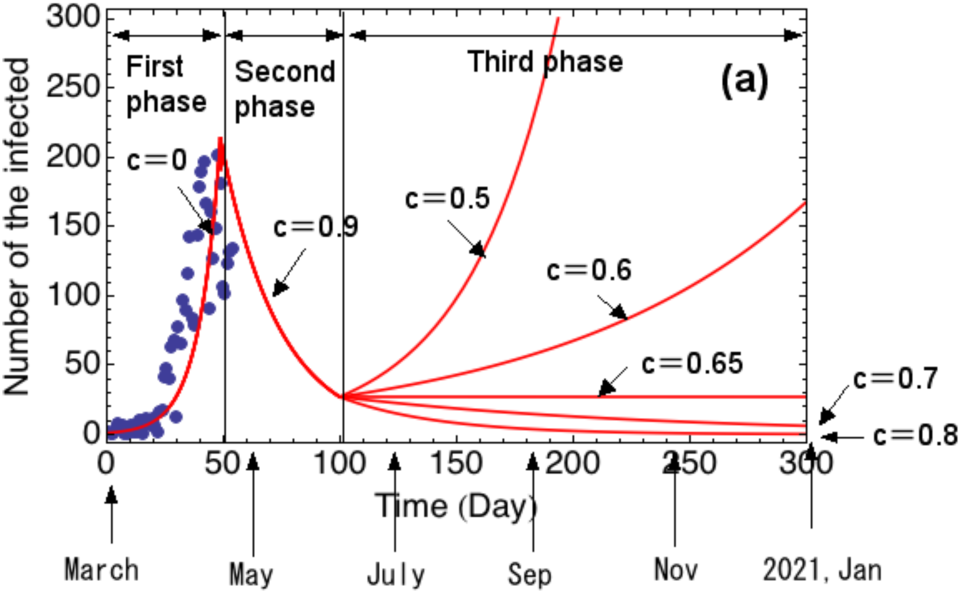
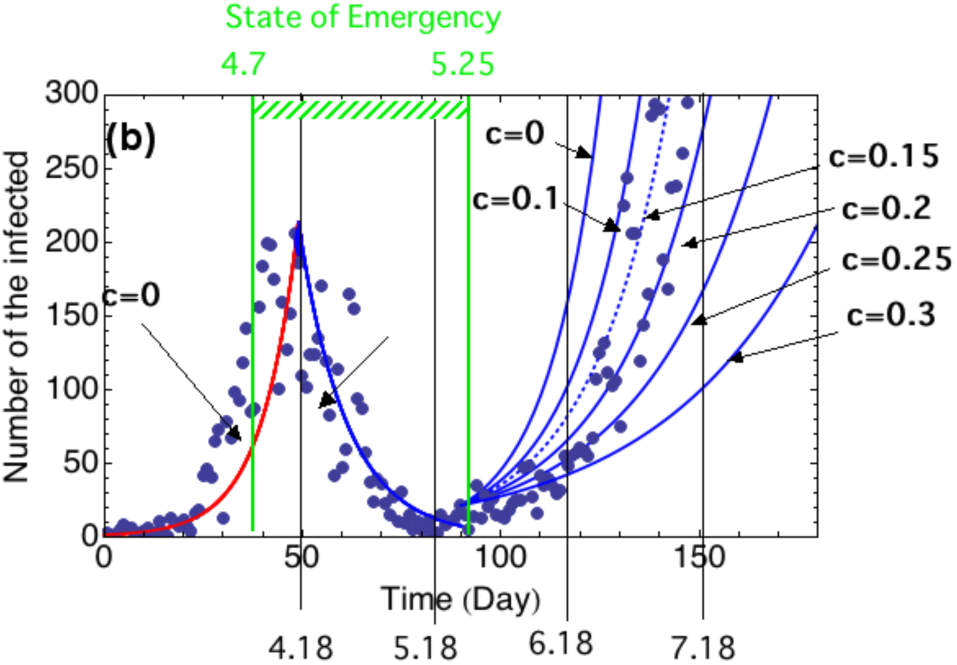
Prediction of the number of the infected as a function of the outing restriction in the third phase until the end of 2020. Each isolation ratio is the same as in Fig.2. (a) Calculated on April, 2020 and (b) calculated on July 11, 2020. Red lines are for γ=0.057 (R_o_=2.9), and blue lines for γ=0.09 (R_o_=1.83).

However, when the outing restriction ratio is given by c=0.65, the number of the infected per days becomes constant. Of course, when c=0.7 or 0.8 are maintained, it will be decreased as seen in Fig. 3-(a). Therefore, to keep the infection number low, the outing restriction ratio should be larger than c=0.65. However, it might be very difficult to maintain such a strong measure from a viewpoint of economy.

When no more restriction is set, the number of infected people rapidly increases as in the first wave. The strong restriction can again suppress the number of the infected. The oscillatory situation can be seen in the report [10]. Therefore, in Tokyo after the state of emergency was lifted up on May 25, 2020, the number of the infected slowly increased as shown in Fig.3-(b). On July 10, 243 persons were confirmed as positive. To estimate the level of outing restriction without the state of emergency, we calculated that the third phase started at Day 90 (21 infected people) and fitted data with curves with c=0 to 0.3 as shown in Fig.3-(b). The isolation ratio of γ=0.09 (the blue line) is assumed in the third phase because the time difference between the confirmed and discharged date from the hospital is almost 11 days after Day 80, which is obtained by the same method as shown in Fig. 1-(b). As the cases of c=0.1 ∼ 0.25 are almost the best fitting curves, the new way of life under the condition without outing restriction, such as mask wearing, hand washing, and social distancing, may be providing this value. We note if the smaller isolation ratio of γ=0.057 is taken, c is further increased as c=0.3 ∼ 0.35. We note that the initial value of the number of patients at Day 90 sensitively determines the subsequent number of the infected people.

## 3. SIR model with outing restriction, vaccination and isolation control

To avoid an outing restriction or lockdown, it is clear that we need to have vaccination. However, to obtain safe vaccine, we have to wait for it. Therefore, we have to manage to keep the number of the infected as low as possible although it is not good for the testing the effect of vaccine. In this section, we propose the comprehensive SIR model to unify the outing restriction, vaccination and isolation together, and show their equivalent relationship.

### 3.1 Outing restriction and vaccination

We show in Fig. 4 how we can consider the SIR epidemic curve when the outing restriction and vaccination are taken into account.

**Fig. 4.**
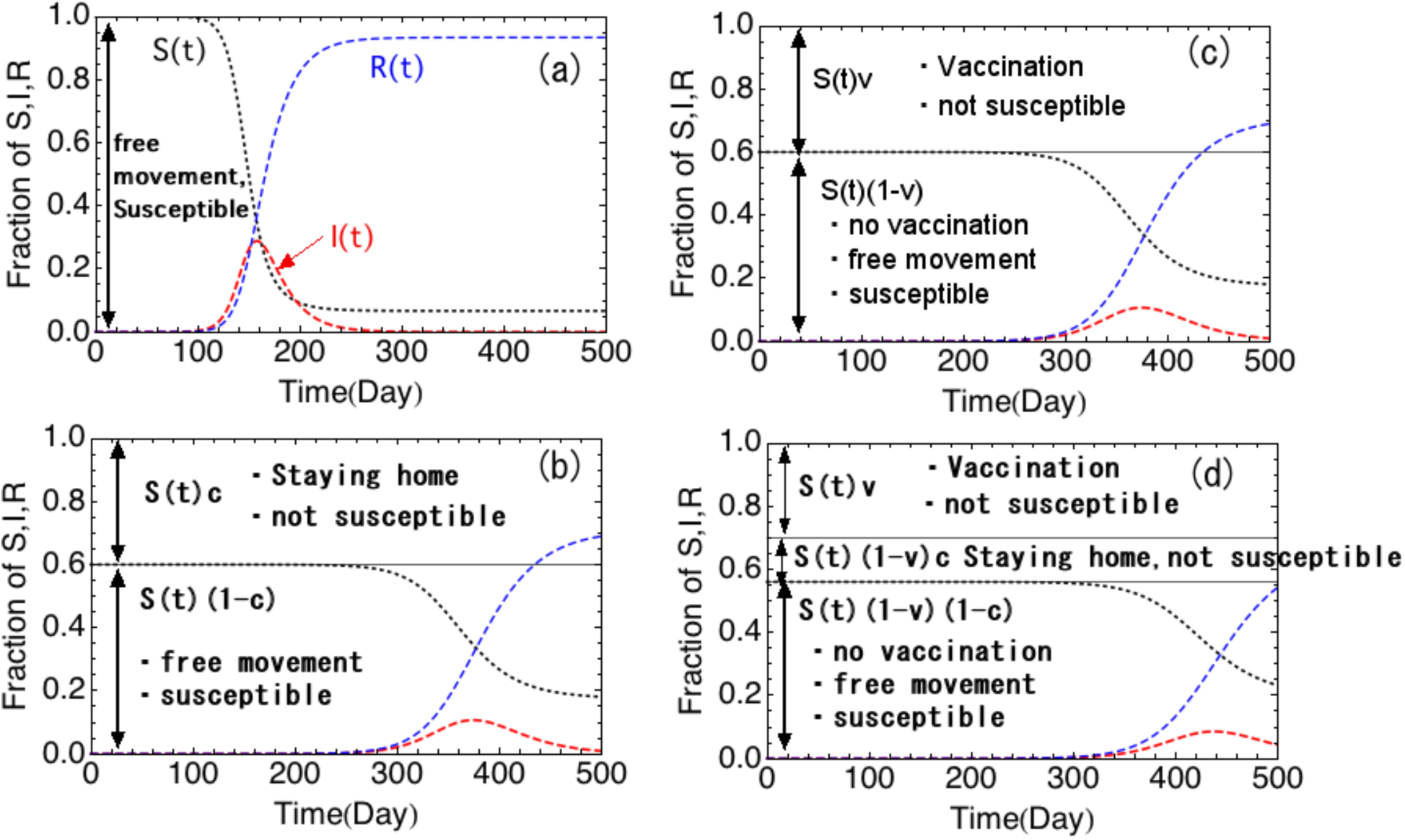
The schematic SIR curves with the outing restriction and vaccination ratios. (a) No outing restriction, (b) Outing restriction alone with c=0.4, (c) Vaccination alone with v=0.4, and (d) vaccination ratio of v=0.3 and outing restriction ratio of c=0.2.

Figure 4-(a) corresponds to no restriction case on the human behavior. The basic reproduction number is R_o_=2.9 as in section 2.1. Figure 4-(b) shows the case of 40 % outing restriction (c=0.4). The number of the susceptible people S(t)(1-c) with free movement is reduced by the outing restriction, and the rest of them S(t)c are staying home. As the horizontal line is given by the initial susceptible number S(0)(1-c), the epidemic curve exists below this line. As the effective reproduction number is R_eff_ = R_o_(1-c) = 1.74 for S(t)∼1 in this case, the peak fraction of the infected I(t) would be delayed and reduced.

When the susceptible people have vaccination with the ratio of v, SIR model equations are given by

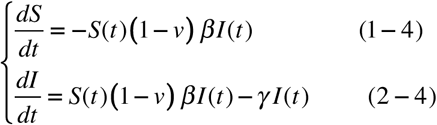

Figure 4-(c) shows this situation with the vaccination ratio of v=0.4.

When the outing restriction c is imposed together with the vaccination ratio v, SIR model equations are modified as

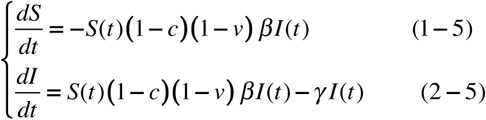

and result is shown in Fig.4-(d) for the vaccination ratio of v=0.3 and the outing restriction ratio of c=0.15. We do see that the infected people appear earlier without any restriction, and later with restrictions.

We show the various SIR curves in Fig. 5 to see the effect of the outing restriction ratio. When the outing restriction ratio c is increased, it is seen that the peak fraction of the infected is reduced and delayed.

**Fig. 5.**
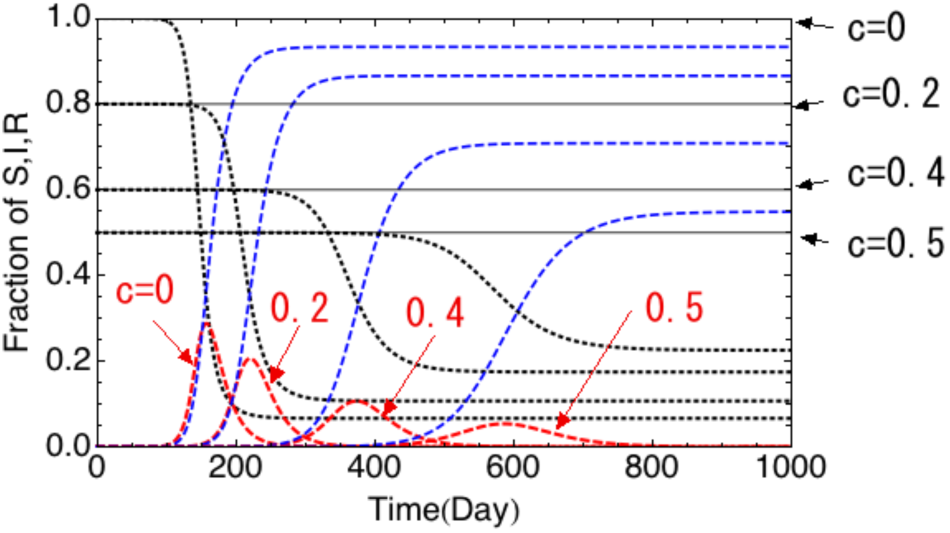
SIR curve depending on the outing restriction ratios c. Black dot line: Susceptible, Red dot line: Infected, Blue dot line: Recovered

The condition to reduce the infected case is given by the effective reproduction number less than 1 as *R*_*eff*_*=S(t)R*_*o*_*(1-c)(1-v)* ≤ *1* when both the outing restriction and vaccination are taken into account. Hence, we have the relationship: *c* ≥ 1-1/{*S(t)R*_*o*_*(1-v)*} which is shown in

Fig 6 for *S(t)*∼1. When the vaccination ratio increases, the outing restriction ratio can be relaxed. Of course this depends on the basic reproduction number as seen in Fig. 6. In the case of large basic reproduction number, the vaccination ratio should be large, because infectious disease is more transmissive. So called “herd immunity” holds for vaccination ratio over ∼50 % without the outing restriction as shown in Fig. 6. However, the herd immunity strategy is quite dangerous because a lot of people are infected and then the death toll will be increased, and also it is not clear yet whether the antibody can last longer or not for COVID-19. We note that if the new way of life with c=0.1∼0.25 is maintained as shown in Fig.3-(b), herd immunity could be established with the vaccination ratio larger than 30 %.

**Fig. 6.**
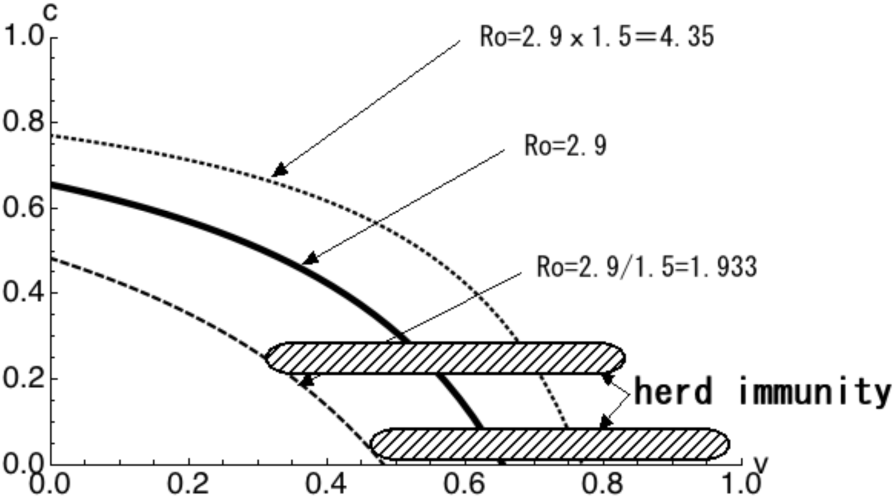
Relationship between the outing restriction ratio c and vaccination ratio v to suppress the infectious disease

At present as we have no effective and safe vaccine, how we can suppress this COVID-19 without outing restriction or lockdown. We seek such a solution based on SIR model.

### 3.2. Isolation ratio γ

The number of the infected people depends on the isolation coefficient γ. This value γ was determined in the first phase where any control measures were not taken yet. Here to know how the number of the infected is increased, we have calculated the first term (solid curve) and the second term (dotted curve) in Eq. (2) for various isolation coefficients γ as shown in Fig. 7. When the isolation coefficient is as small as γ = 0.04, the peak value of the infected people βS(t)I(t) is much larger than the one of the second term γI(t). We see that the second term is delayed from the first term. Therefore, the infected people are increased. For example, γ = 0.057 has been used in the analysis for Tokyo as shown in Figs. 1, 2 and 3.

**Fig. 7.**
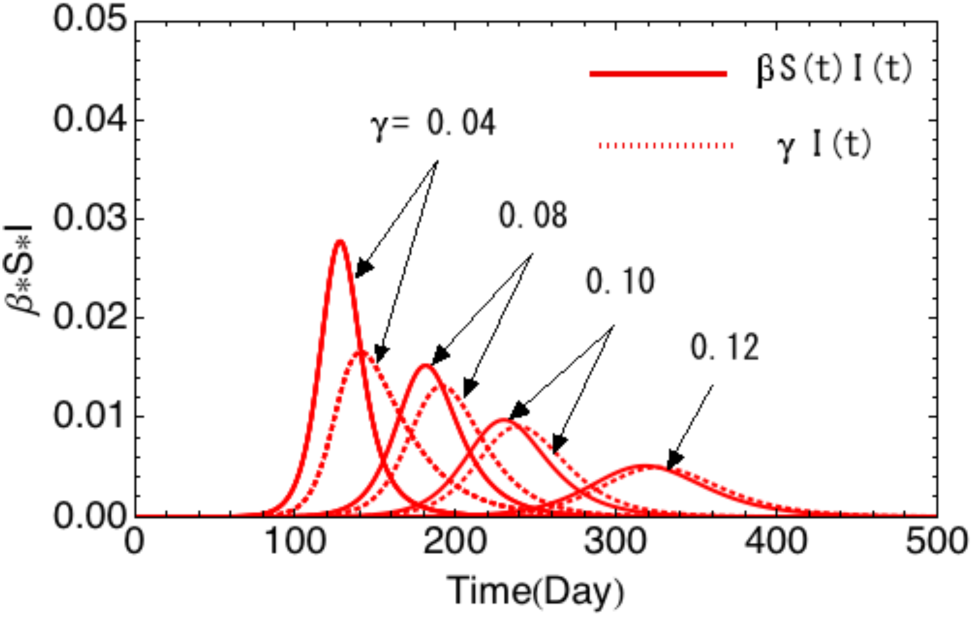
The time dependence of the first term and second term in Eq.(2) for various isolation ratios

On the other hand, for the larger isolation coefficient as γ= 0.12, the difference of their peaks is small and their time delay becomes shorter with the isolation ratio. In other word, the first term and second term become comparable. This means that the number of the infected is not increased if the isolation is taken as soon as the case is confirmed. This is also interpreted as follows. As the basic reproduction number is given by R_o_ = β/γ, the large isolation coefficient γ provides the smaller R_o_, reducing the number of infected people.

### 3.3. Equivalence between outing restriction and isolation in SIR model

Outing restriction (or lockdown) causes the large economical loss. Therefore, we should consider how this epidemic could be overcome without vaccine and outing restriction.

When the outing restriction ratio of c is imposed, the effective reproduction number is given from Eq. (2-3) as *R*_*eff*_*=βS(t)(1-c)/γ*. On the other hand, when the excess isolation q takes place (no infected person is isolated by testing error), we can use *γ(1+q)* as the isolation term coefficient. The effective reproduction number is given by

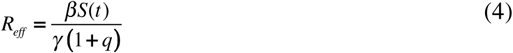

in this case. This leads to the following linear approximation in the case of q <<1 as

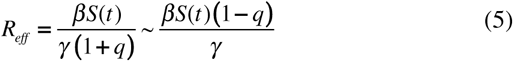

This is exactly the same expression with the outing restriction ratio of q. We have thus found that there is the equivalence between outing restriction (lockdown) and isolation in Eq. (2) in SIR model. However, this holds only for the small value of q less than 1.

If we introduce the isolation term as

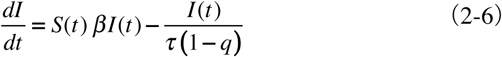

it can be used for the large value close to q = 1. This provides the effective reproduction number *R*_*eff*_*=βS(t)/γ* = *βS(t)τ (1-q)= S(t)R*_*o*_ *(1-q)* with *R*_*o*_= *βτ*.

The large q value provides the smaller effective reproduction number. This situation is shown in Fig. 8. During this recovering time, patient is infectious but not actually isolated from the susceptible space so far in the traditional SIR model. By introduction of the isolation time control parameter q, we can artificially control this infectious time.

**Fig. 8.**
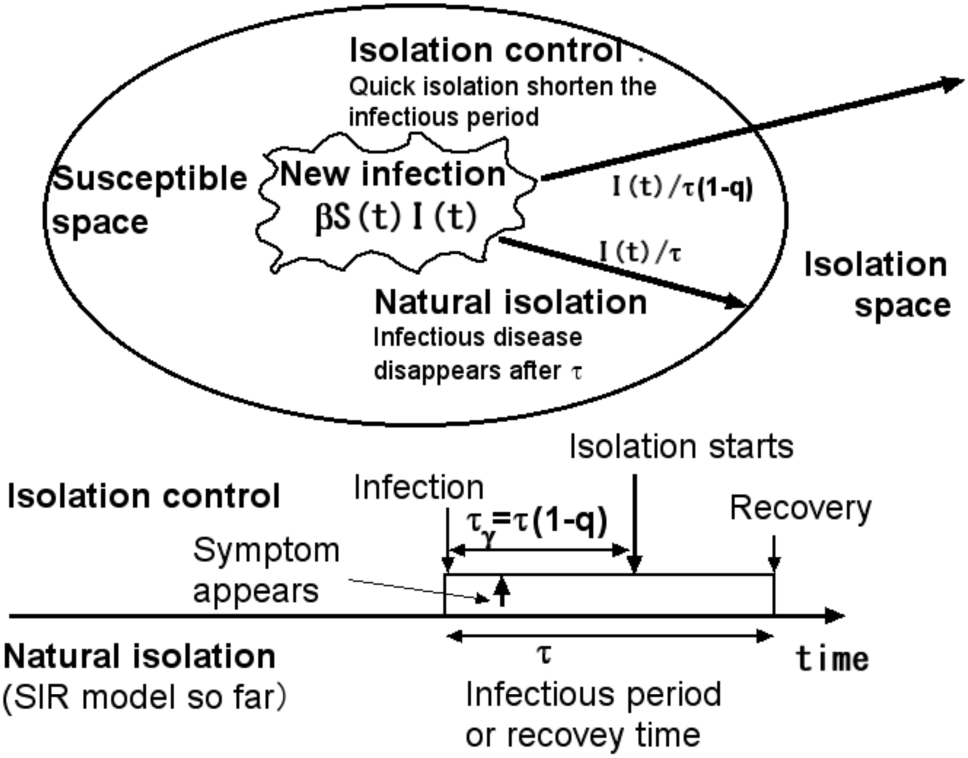
Concept of natural isolation in SIR model and isolation time control proposed in this work.

Isolation control has the same effect as the outing restriction. To confirm this, we calculated the second phase using the isolation time control by changing q from 0.2 to 0.8 as shown in Fig.9. The number of the infected actually decreases with the isolation time control parameter q as the outing restriction case. The required isolation times τ_γ_ =τ(1-q) are also shown in the parenthesis for each q. It is seen that 3 to 5 days are necessary to reduce the number of the infected to the small value. The horizontal line is obtained when *R*_*o*_*(1-q)*=1 is satisfied, namely q=0.65 for *R*_*o*_=2.9, which agrees with the numerical one. If the isolation time control parameter q is larger this value, the number of the infected decreases.

The required isolation time can be longer when the outing restriction is imposed at the same time as will be seen in the next section. The required isolation time thus obtained becomes quite reasonable. If isolation can be done as soon as the infected is identified, it reduces the infection.

### 3.4. Combination of the mild lock down and isolation time control

When the outing restriction is imposed at the same time, isolation control is relaxed. The isolation time can be longer than that with isolation control alone.

Equation (2-6) can be written in this case as

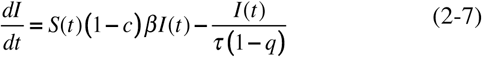

and the effective reproduction number is given by *R*_*eff*_*=βS(t)/γ* = *S(t)R*_*o*_*(1-c) (1-q)*. This means that the outing restriction is equivalent to the isolation control. For example, as the outing restriction ratio of c = 0.3 and isolation time control parameter of q = 0.7 provides (1-c)(1-q)= 0.7×0.3∼0.21, which is almost the same as the outing restriction ratio of c = 0.8.

Long term prediction until the end of 2020, corresponding to Fig. 3, is conducted for various isolation time control parameters of q=0.5 to 0.7. Results are shown in Fig.10 in the third phase, how the number of the infected is controlled by the isolation time control parameter together with the outing restriction of c=0.3, corresponding to the parameters in the new way of life as shown in Fig.3-(b).

For q=0.55, the number of the infected is reduced to 3 persons per day at the end of 2020. For the case of q=0.7, the number of the infected is 0.6 person after 50 days. Thus we have found that the longer isolation time is permitted when outing restriction is imposed at the same time, which could be called the combined control.

The condition of the flat curve in the third phase given by *S(t)R*_*o*_*(1-c)(1-q) =1* provides *q=1-1/{R*_*o*_*(1-c)}* = 0.50 for *S(t)*∼1, *R*_*o*_=2.9 and c=0.3 as shown in Fig.10.

To compare the case without the outing restriction in the third phase, the same condition is used except for c=0. To suppress the infectious disease it is necessary to isolate the infected people within 5.2 days (q=0.7) without the outing restriction as shown in Fig. 11. However, with the outing restriction ratio of c=0.3, 7.8 days (q=0.55) is enough to suppress (Fig. 10). Thus we have found that it is easier to suppress the infectious disease by isolation control if the mild outing restriction is imposed at the same time.

**Fig. 9.**
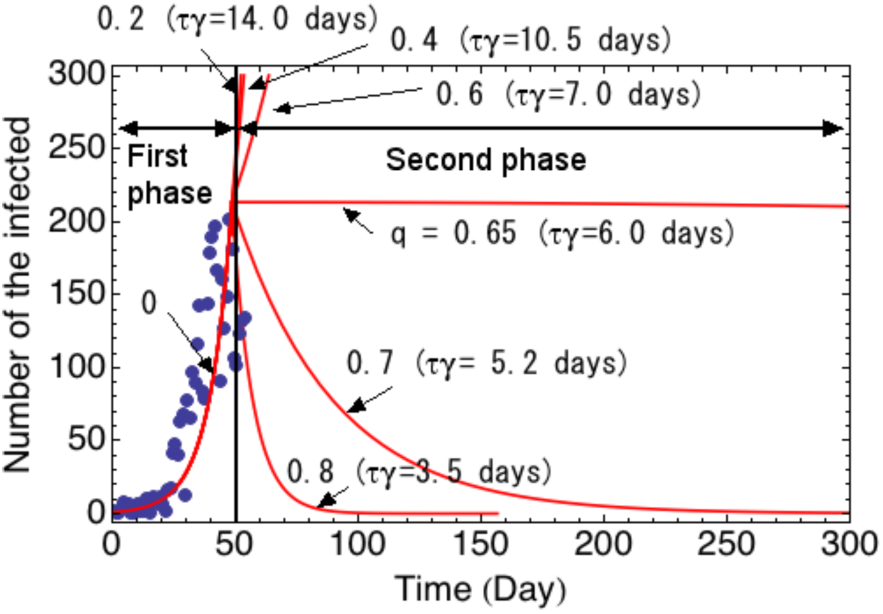
Temporal evolution of the number of the infected for various isolation time control parameters q without any outing restriction c=0. (τ_γ_= τ (1-q) and τ =17.54 days). Red lines are for γ=0.057 (R_o_=2.9).

**Fig. 10.**
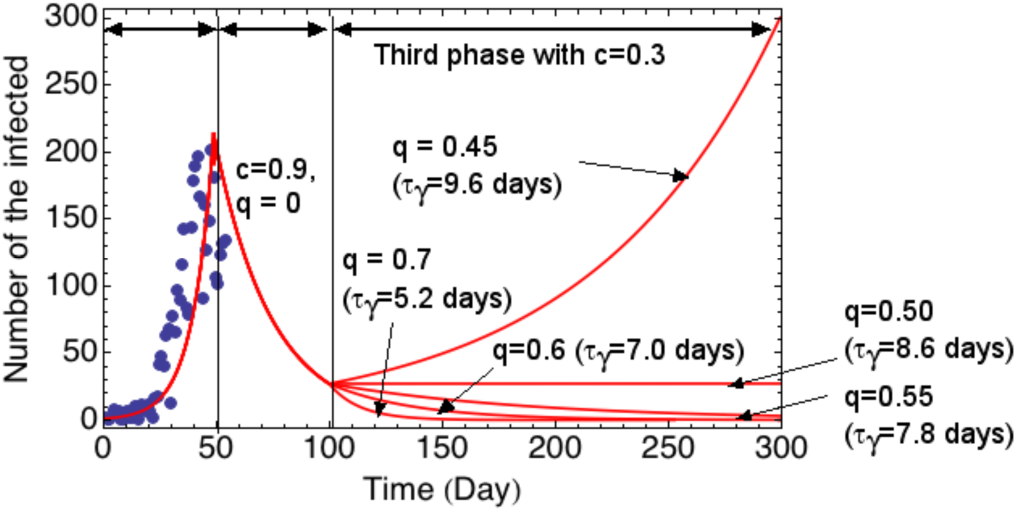
Dependencies of the number of the infected on isolation time control parameters q with the outing restriction of c=0.3. (τ_γ_= τ (1-q) and τ =17.54 days).

**Fig. 11.**
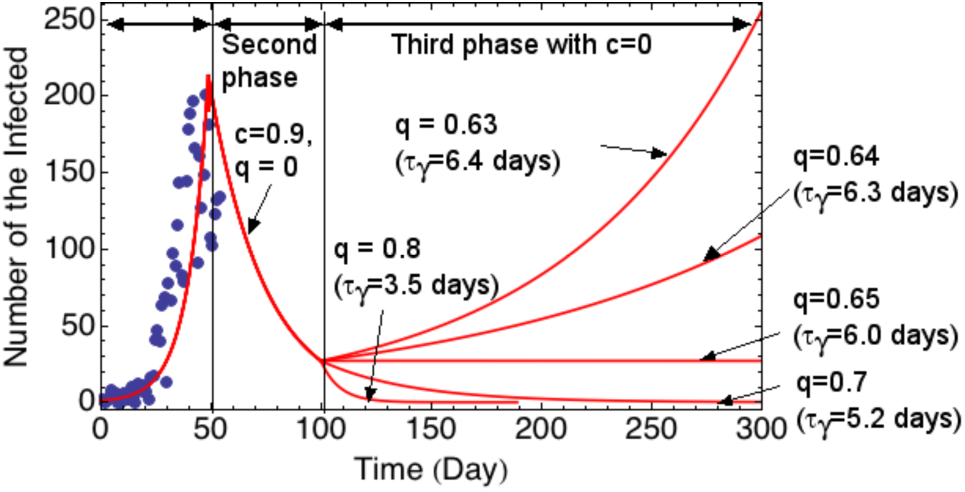
Dependencies of the number of the infected on isolation time control parameters without the outing restriction c=0. (τ_γ_= τ (1-q) and τ =17.54 days).

When peoples have vaccination with outing restriction, Eq. (2-7) is finally given by

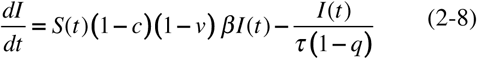

and the effective reproduction number is given by *R*_*eff*_*= S(t)R*_*o*_*(1-c) (1-v) (1-q)*. The vaccination ratio of v = 0.3 provides the same effect with the outing restriction ratio of c=0.3. Therefore, by vaccination ratio of v=0.3, it is possible to suppress this infection disease within 2020 by the isolation time control parameter with q > 0.55 (isolation within 7 days) as shown in Fig.10 without outing restriction. Thus, not to affect the daily life and social economy, this isolation time control strategy is a hopeful solution.

## 4. Discussions and summary

We have noticed the interesting papers on the effectiveness of isolation and quarantine to overcome infectious disease [11]. We have used the word “isolation” as used in the reference [12]. Accurate data on viral load with respect to the time from infection as shown in the reference [13][14]. could improve the estimation proposed in this work. Because only average time has been used in this work. However, such work is beyond this paper.

Big problem in this COVID-19 is that there are a lot of asymptomatic but infectious cases [15][16]. Therefore, active and massive testing is so important for quick isolation of the identified person. Such technology has been under active development. For example, recently the new method is invented by TakaraBio in USA to allow faster and larger scale PCR testing, where 5,000 tests can be done in about two hours [17]. As the National Institute of Health is aiming at the 6 million PCR tests per day, such aggressive research plan is promising to suppress this pandemic [18]. Recently, innovative technique such as vocal biomarkers is proposed by MIT Lincoln Laboratory to identify the asymptomatic people with COVID-19 positive [19]. Vocal change before and after infection could be detected by processing speech recording. In addition, contact-tracing technique using communication tool is also useful [13].

There is a well known method to survey the COVID-19 spread in the country with the small capacity of the PCR testing resources. As swage surveillance successfully used for monitoring Polio [20], it might be an effective method to detect the COVID-19 infected peoples in the restricted area within the swage network [21]-[24]. As it can find the COVID-19 positive without time delay, the actual COVID-19 spread in that area is provided. If the peoples living in such area are tested by group PCR testing, it can reduce the PCR testing resources and shorten the isolation time. Therefore, such swage water monitoring system should be constructed for monitoring the spread of COVID-19 infection to terminate this pandemic. It might be also useful for future unknown infectious diseases.

Research on the detector dog is also very interesting to identify the COVID-19 patients. As reported in the papers [25][26], the trained dog can distinguish the COVID-19 patient by their scent. Average detection rate is reported to be 94 % [25]. Therefore, this method might be very useful in mass gathering events and airport where quick and massive PCR testing is required. Various new ideas and wisdom as discussed above should be taken up seriously to overcome this pandemic.

To know the truth of infectious disease, we have studied the characteristics of SIR model equation in detail. Based on the ^4^He ash removal study in a D-T nuclear fusion [1], we have found that the time in the isolation term can be interpreted as the natural isolation time, which means that the isolation is not conducted in a controlled manner in a traditional SIR model. Therefore, we introduce the isolation time control parameter q to control the isolation time. We have finally found that the outing restriction ratio c, the vaccination rate v, and the isolation time control parameter q are equivalent with respect to the basic reproduction number. This means that isolation control has the same function of vaccination or the outing restriction. In an actual situation especially after the first wave when the number of patient decreases, massive PCR testing etc should be conducted and then infected people especially without symptom should be identified and isolated as soon as possible. We have to construct such an advanced social system with great care about human rights. Without such system, the economic activity cannot be maintained unless a safe vaccine is not supplied.

In this study, the required isolation time is 3 ∼ 5 days without any outing restriction, but it can be longer as 5 ∼ 8 days with the outing restriction of c=0.3 to suppress this COVID-19. We note that this value is obtained by the new way of life as shown in Fig.3-(b). Although this isolation time depends on the basic reproduction number, the required isolation time obtained in this study is quite realistic and could be accomplished. Above estimated time could be also justified by the fact that the median duration of viral shedding in the asymptomatic group is 19 days [16]. This value is quite similar to our employed value of infectious period or recovery time τ =17.54 days. This isolation time control concept may be able to explain the success in Taiwan, New Zealand and Mainland China to suppress COVID-19 disease.

In conclusion, with reliable, simple and massive testing method, if isolation can be done in a short time after case identification, this pandemic could be overcome without the strong outing restriction such as lockdown before vaccine is completed.

Medical research institutes and pharmaceutical companies are actively developing the vaccine to curtail this pandemic. In contrast, only the central and local governments can do wide, repeated and quick COVID-19 testing and quick isolation. The bureaucratic system such as the public health center and the ministry of health can play an important role to manage the isolation time control together with various laws. Upon considering this simple theory proposed here, we believe government should be definitely a main player to terminate this infectious disease.

## Data Availability

The following two datasets have been used.

https://oku.edu.mie-u.ac.jp/~okumura/python/COVID-19.html

https://toyokeizai.net/sp/visual/tko/covid19/

## Conflict of interest

None declared

## Author statement

O.Mitarai: Conceptualization, Methdology, Software, Writing, Original draft

N.Yanagi: Conceptualization, Supervision, Writing

## Funding

None

## Acknowledgement

Publication of this work is supported under the auspices of the National Institute for Fusion Science (NIFS) Collaborative Research Program.

## Appendix. Analogy between nuclear fusion research and SIR model

In a deuterium and tritium nuclear fusion reactions, the following reaction takes place.

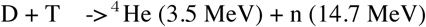

Here high energy alpha particles (^4^He particle) should be removed from the D-T plasmas after losing their energy to the surrounding D-T plasmas. This ^4^He particle can be expressed by the particle balance equation given by [Eq.(3) in the Ref. [1]]

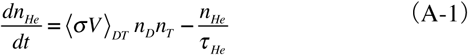

The first term shows the ^4^He production term by D-T nuclear reactions, where n_D_ is the deuterium, density, n_T_ is the tritium density, <σV>_DT_ is the D-T fusion rate, σ is the cross section of the D-T fusion and V is the relative velocity between D and T particles. This term is also analogous to the first term in Eq. (2) in the SIR model, which expresses the collision (close contact) between the susceptible and infective persons.

The second term expresses the ^4^He particle escaping from the plasma. Escaping time is τ_He_ in an average as shown in Fig.A. This term is exactly the same as the isolation term in SIR model equation (2). This can be interpreted as that the infected person goes out to the isolation space after τ_γ_. During this infectious time the infected persons moves freely. The infected people is not isolated immediately after infection as this He ash.

**Fig. A.**
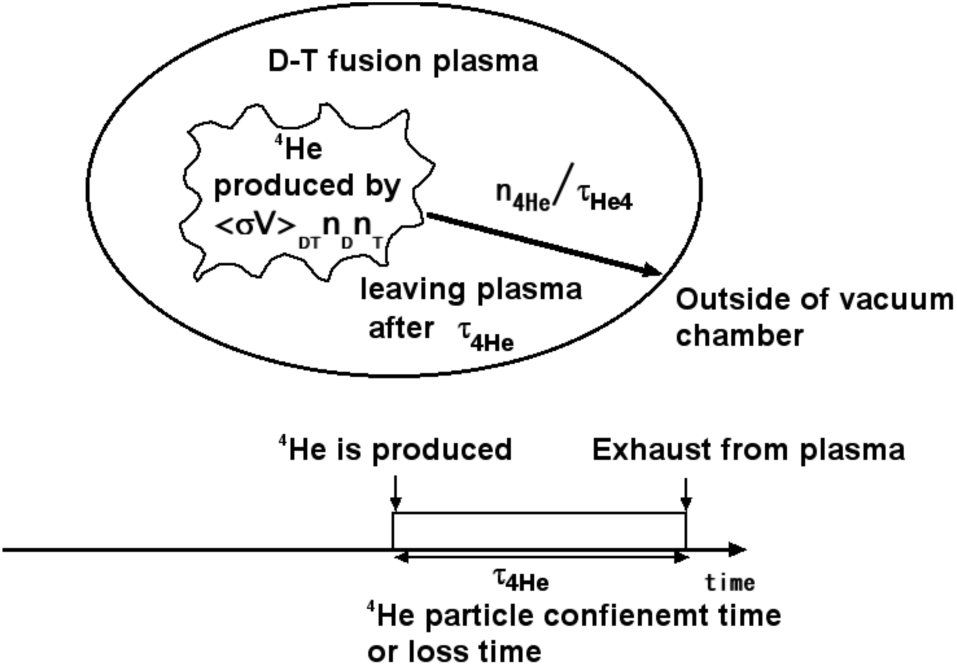
He ash exhaust in a D-T fusion plasma

We have long been studying how we can exhaust these ^4^He ash particles quickly from the fusion plasma such as in tokamak and helical reactors. We have noticed that especially in an advanced fusion reactor such as neutron lean fusion reactor, exhaust of ash particles are crucially important [27]. The particle confinement time in the second term should be as short as possible to build a fusion reactor. This fusion concept is the quite similar to the epidemic strategy to maintain the economic activity by isolating the infected people quickly.

## Notes

### Competing Interest Statement

The authors have declared no competing interest.

### Author Declarations

(1)Institute for Advanced Fusion & Physics Education, 2-14-8 Tokuou, Kitaku, Kumamoto, 861-5525, Japan (2)National Institute for Fusion Science, 322-6 Oroshi-cho, Toki, Gifu, 509-5292, Japan

